# Circles of Care: a geospatial analysis of computed tomography (CT) imaging access and socioeconomic vulnerability in Houston, Texas

**DOI:** 10.64898/2026.04.28.26351996

**Authors:** Daniel Martinez Ortiz

## Abstract

**Background:** Computed tomography (CT) is a cornerstone of timely diagnosis for stroke, trauma, and oncologic conditions, and delays in access are associated with worsened outcomes. Although Houston, Texas, is home to one of the world’s largest medical complexes, the geographic distribution of CT imaging infrastructure has not been systematically examined against neighborhood-level measures of socioeconomic vulnerability.

**Methods:** We conducted a cross-sectional geospatial analysis of CT imaging facilities across the Greater Houston metropolitan area. Facility locations — including hospital-based scanners, independent imaging centers, and freestanding emergency facilities — were compiled from publicly available imaging directories, Texas Department of State Health Services (DSHS) facility listings, Centers for Medicare & Medicaid Services (CMS) provider data, and CT location data contributed by MD Anderson Cancer Center. Census tract–level indicators (median household income, percent uninsured, poverty rate) were obtained from the U.S. Census Bureau American Community Survey. Facility locations were geocoded and overlaid on census-tract choropleths in ArcGIS Online and ArcGIS StoryMaps to identify tracts with elevated socioeconomic vulnerability and limited proximity to CT infrastructure.

**Results:** CT imaging facilities were markedly clustered in the central urban core and in higher-income corridors, with hospital-based and independent scanners concentrated in census tracts with lower poverty rates, higher median household income (>$119,300), and higher insurance coverage. Conversely, peripheral and southeastern tracts with elevated poverty (>24%), median household income below $37,800, and uninsured rates exceeding 16% contained comparatively sparse CT infrastructure, generating spatial “gaps” in advanced diagnostic capacity. The pattern persisted across facility type: freestanding emergency and independent imaging centers did not meaningfully compensate for the undersupply of hospital-based scanners in vulnerable communities.

**Conclusions:** In Houston, the spatial distribution of CT imaging resources mirrors rather than offsets underlying socioeconomic inequality. Neighborhoods with higher poverty and uninsured rates face compounded barriers of distance and coverage. Citywide spatial analysis renders these inequities visible in ways individual clinical encounters cannot, and supports equity-informed health-system planning, targeted investment in underserved catchments, and policies linking imaging-capacity expansion to measurable community need.

## Background

Computed tomography (CT) is a foundational modality in contemporary clinical medicine, providing rapid cross-sectional imaging that underpins the evaluation of acute stroke, traumatic injury, oncologic staging, pulmonary embolism, and a wide range of emergency and time-sensitive conditions [1, 2]. Because clinical decisions in many of these conditions depend on imaging results obtained within narrow diagnostic windows, delays in access to CT have been repeatedly associated with worsened outcomes, higher downstream utilization, and missed opportunities for early intervention [2, 3]. Consequently, the physical and organizational distribution of CT capacity across a health system has real consequences for population health and for equity in care [4].

The geographic dimension of access to health services has been extensively studied under the rubric of spatial accessibility, including two-step floating catchment area (2SFCA) methods and their extensions [5, 6]. A consistent finding across U.S. and international settings is that advanced diagnostic infrastructure tends to concentrate in urban cores and affluent suburbs, while peri-urban and rural communities — and, within cities, lower-income and higher-uninsured neighborhoods — often face disproportionate travel burdens and longer appointment wait times [5–7]. These structural patterns compound patient-level barriers such as lack of insurance, limited transportation, and time off work, and they contribute to disparities in diagnosis and outcomes that are increasingly visible in public-health surveillance systems [7, 8].

Houston, Texas is a particularly informative setting in which to examine these questions. Greater Houston is home to the Texas Medical Center — one of the largest concentrations of hospitals, imaging centers, and specialty services in the world — yet substantial socioeconomic heterogeneity exists across its census tracts [9]. Prior analyses have documented wide disparities in insurance coverage, primary-care access and chronic-disease burden across the city [9], but comparatively little work has rigorously characterized how advanced diagnostic infrastructure specifically — including CT — is distributed relative to neighborhood-level measures of socioeconomic vulnerability. Public-health planning frequently assumes that the sheer density of clinical resources in Houston provides adequate access; a spatially explicit analysis is needed to test that assumption.

This study addresses that gap. Using publicly available data, a facility roster contributed by MD Anderson Cancer Center, and the American Community Survey (ACS), we conducted a cross-sectional geospatial analysis of CT imaging facilities across Greater Houston. Facility locations were overlaid on census tract–level choropleths of poverty rate, percent uninsured, and median household income to assess whether the distribution of advanced diagnostic infrastructure aligns with or diverges from community-level indicators of need. We hypothesized that CT imaging capacity would be concentrated in higher-income, better-insured tracts in the urban core and undersupplied in peripheral and economically disadvantaged tracts, producing geographic “gaps” in access that are invisible at the level of any single clinical encounter but visible at citywide scale.

## Methods

### Study design and setting

We conducted a cross-sectional, descriptive geospatial analysis of CT imaging facilities located within the Greater Houston metropolitan area (Harris County and adjacent counties, including Fort Bend, Brazoria, Galveston, Montgomery, Liberty and Waller). Facility and census-tract indicators reflect the most recent publicly available vintages at the time of analysis (2024–2025). No individual-level patient data were used; the unit of analysis was the census tract.

### Data sources

CT imaging facility locations were compiled from four complementary sources: (i) publicly available imaging-center directories and state licensure listings maintained by the Texas Department of State Health Services (DSHS) [15]; (ii) facility-level information from Centers for Medicare & Medicaid Services (CMS) public-use provider files [14]; (iii) institutional websites of hospital systems operating in Greater Houston; and (iv) CT imaging location data contributed by The University of Texas MD Anderson Cancer Center. Facilities were classified by type into hospital-based scanners, independent imaging centers, and freestanding emergency facilities. Duplicate facility records were resolved by manual review.

Census tract–level socioeconomic indicators were obtained from the U.S. Census Bureau American Community Survey (ACS) 5-year estimates [13]. The indicators examined were median household income, percentage of residents without health insurance (among residents aged 18–64 years), poverty rate (population below the federal poverty line), and total population and population density. Additional geographic reference layers (roads, waterways, place labels and Houston city limits) were drawn from OpenStreetMap, Esri Living Atlas, City of Houston GIS, and the ArcGIS World Imagery basemap [16].

### Geocoding and spatial analysis

Facility addresses were geocoded using the ArcGIS Online World Geocoding Service. Each geocoded location was validated by visual inspection against street-level imagery; records with match scores below established thresholds were corrected manually against institutional websites. ACS tabular indicators were joined to the 2020 Census tract boundary file for the study area. Choropleth maps of poverty rate, uninsured rate, and median household income were generated at the census-tract level, and CT facility points were overlaid to produce the visualizations presented in Figures 1–4. Spatial overlay analysis was then used to characterize concordance between facility density and indicators of community need. For the summary counts reported in Table 1, facilities falling within tracts meeting each vulnerability threshold (poverty rate >24%, uninsured rate >16%, median household income <$37,800) were enumerated by visual overlay of the geocoded facility points on the filtered tract choropleth; counts are approximate to ±5 facilities and are intended as descriptive, not inferential, summaries of the spatial pattern.

**Table 1.** CT imaging facility counts by census-tract socioeconomic vulnerability threshold, Greater Houston, 2024–2025.

| Vulnerability indicator | Threshold | Facilities in threshold tracts (n) | % of total (n = 408) |
| --- | --- | --- | --- |
| Poverty rate | >24% | 45 | 11.0% |
| Uninsured rate | >16% | 20 | 4.9% |
| Median household income | <\$37,800 | 25 | 6.1% |

*Facility counts within vulnerability thresholds were determined by visual overlay of geocoded facility points on filtered census-tract choropleths in ArcGIS Online; counts are approximate to $\pm 5$ facilities. Threshold tracts are defined as census tracts meeting or exceeding the indicated cutoff for each socioeconomic indicator.*

**Figure 1.**
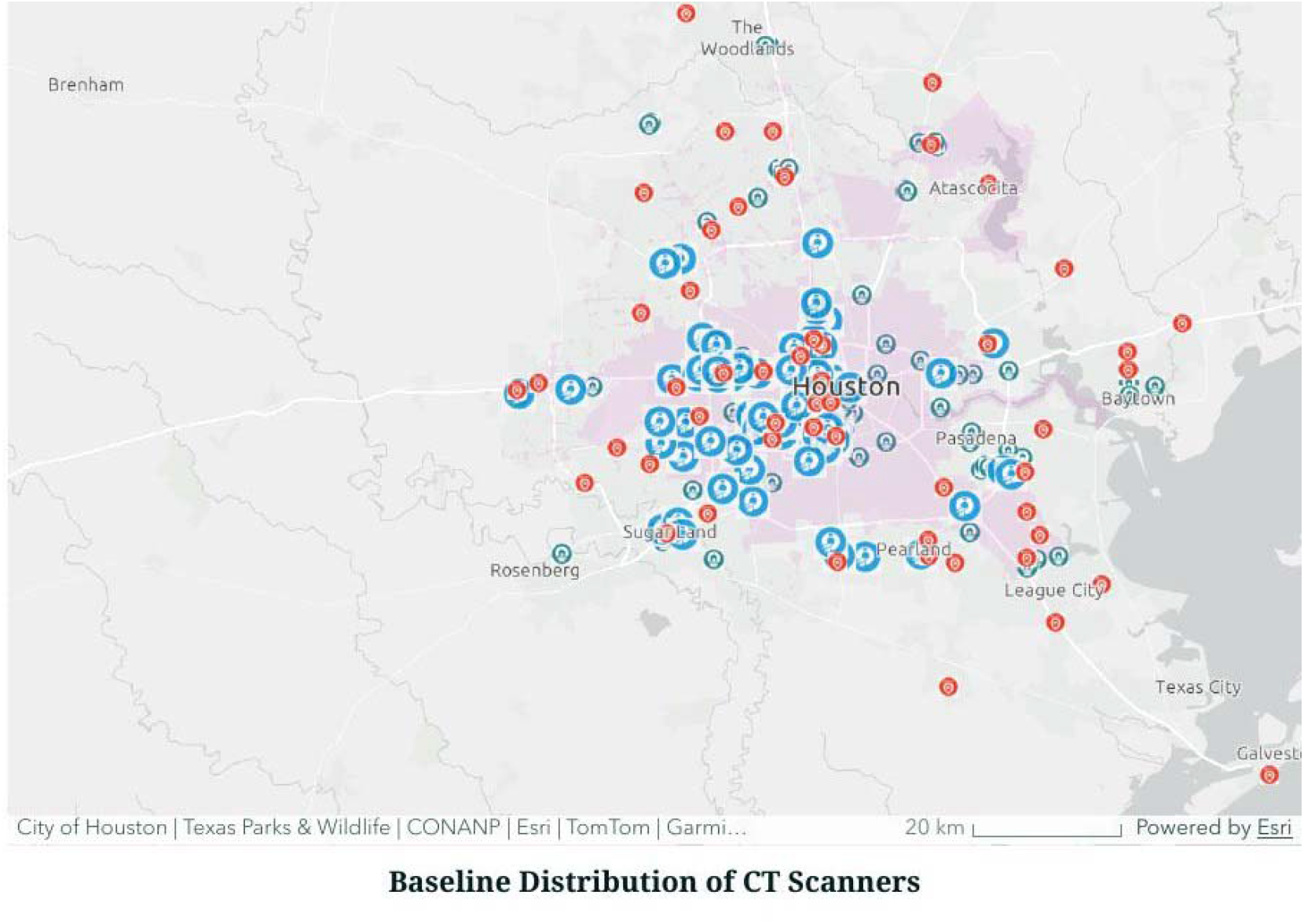
Baseline distribution of CT imaging facilities across Greater Houston, including hospital-based scanners, independent imaging centers, and freestanding emergency facilities. The pink boundary denotes the Houston city limits.

**Figure 2.**
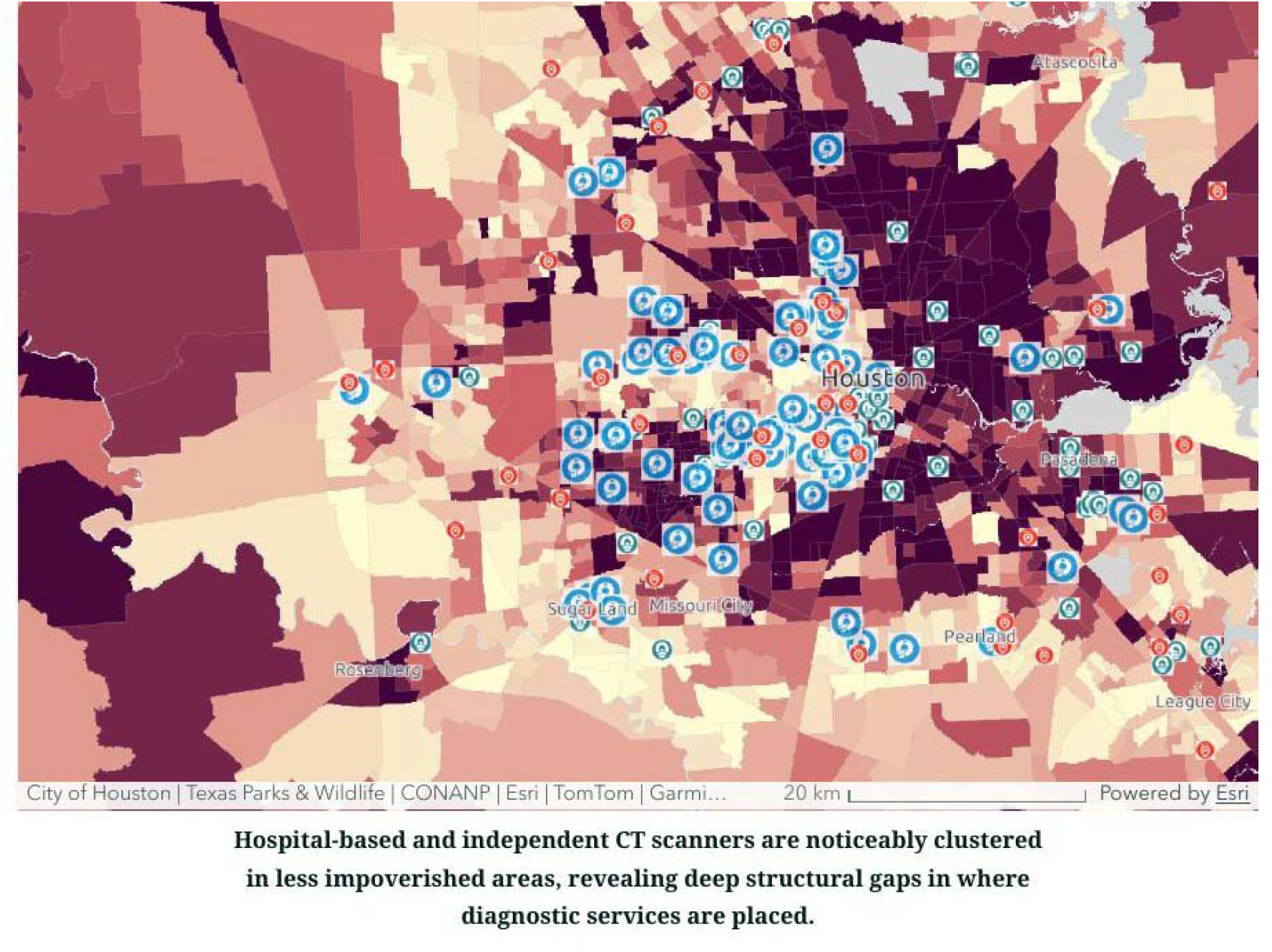
CT facility locations overlaid on a census-tract choropleth of poverty rate (lighter shading = lower poverty; darker shading = higher poverty). Hospital-based and independent CT scanners are noticeably clustered in less impoverished areas, revealing structural gaps in where advanced diagnostic services are placed.

**Figure 3.**
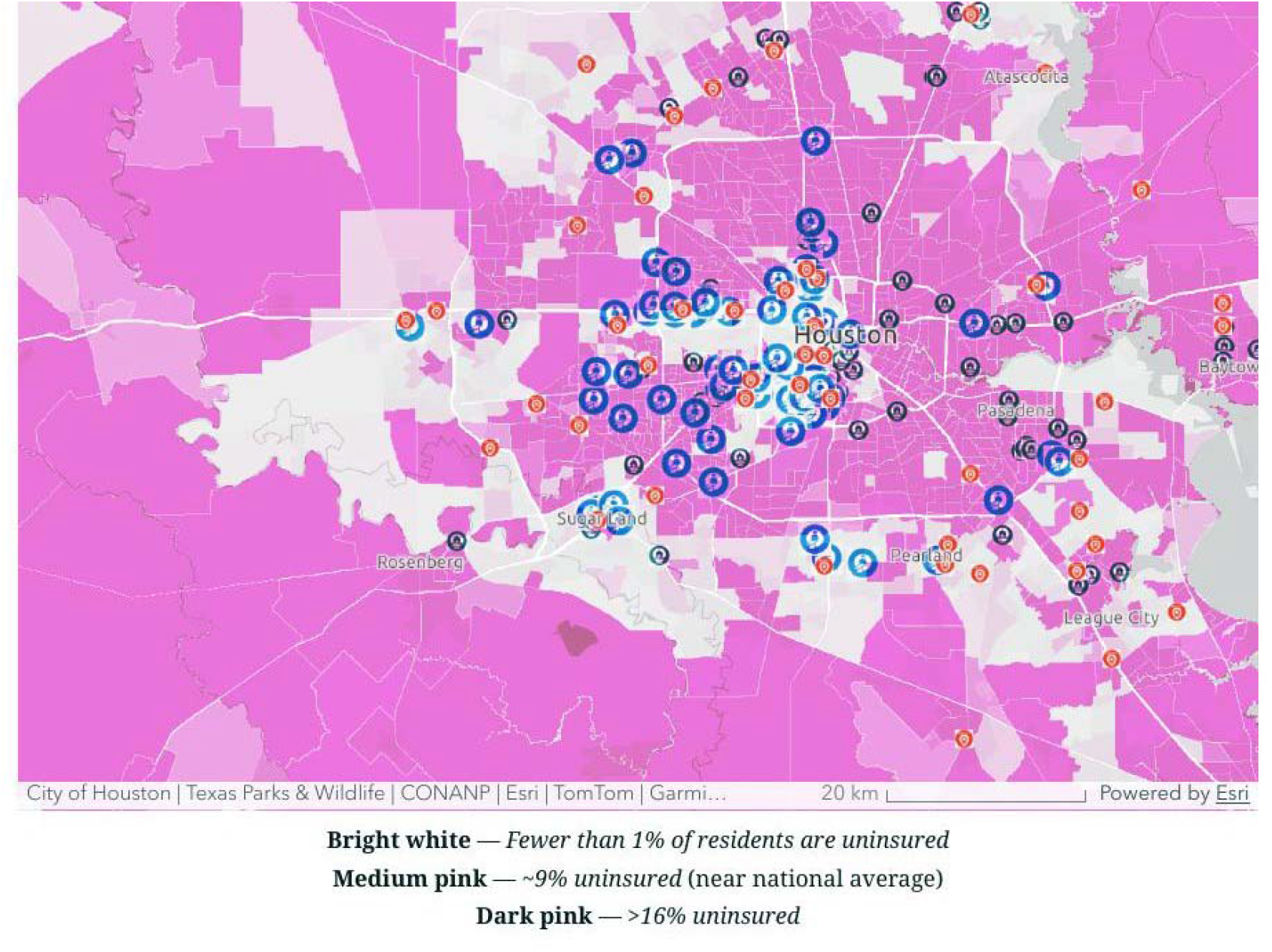
CT facility locations overlaid on a census-tract choropleth of uninsured rate. Bright white: <1% uninsured; medium pink: ∼9% uninsured (near national average); dark pink: >16% uninsured. Tracts with higher uninsured rates on the urban periphery contain markedly fewer CT imaging facilities than the central, better-insured core.

**Figure 4.**
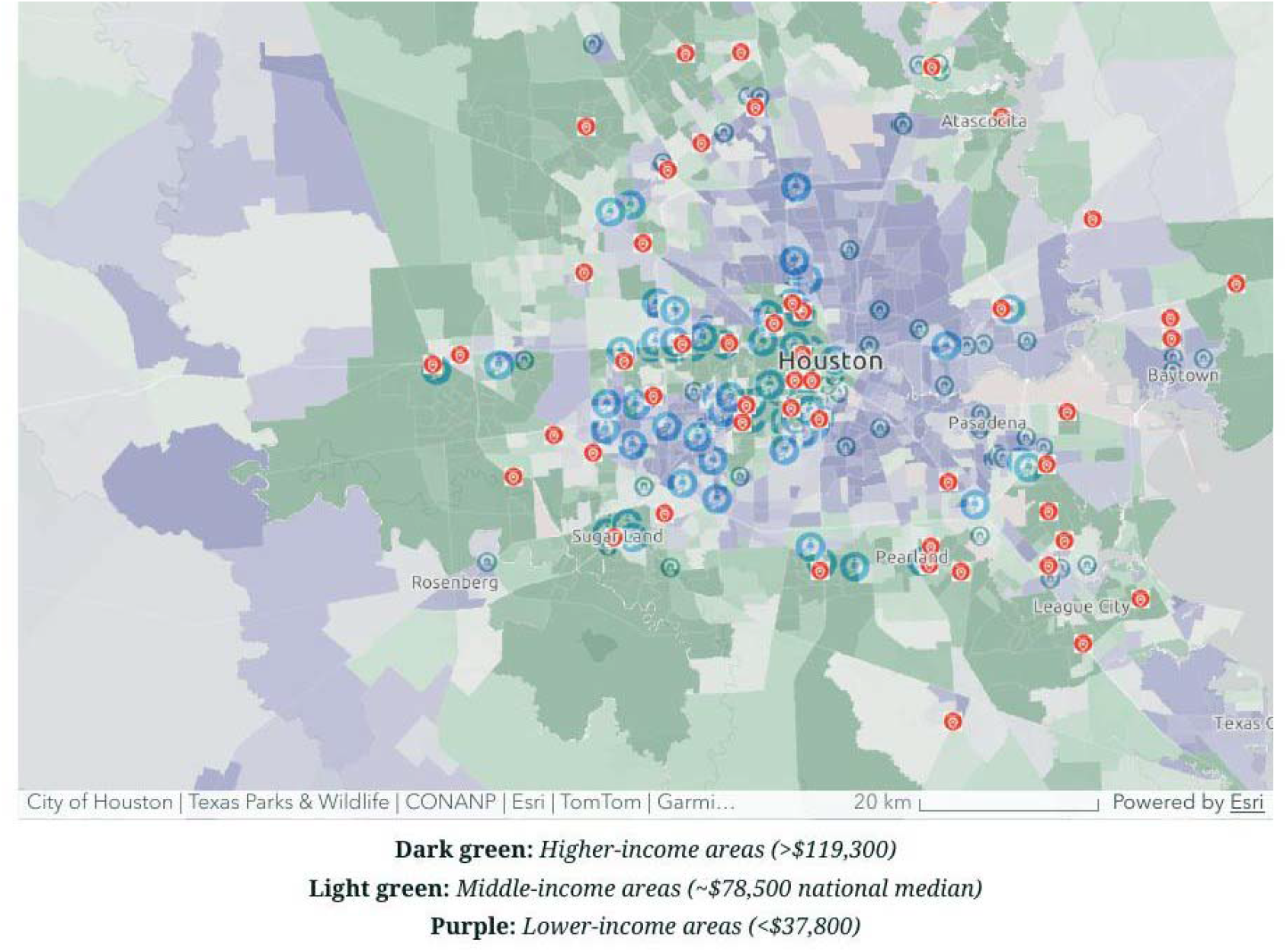
CT facility locations overlaid on a census-tract choropleth of median household income. Dark green: higher-income tracts (>$119,300); light green: middle-income tracts (∼$78,500, near the national median); purple: lower-income tracts (<$37,800). A clear pattern emerges in which CT scanners cluster in higher-income zones while lower-income neighborhoods exhibit fewer nearby diagnostic resources.

### Software

All mapping and geospatial analysis were performed in ArcGIS Online and ArcGIS StoryMaps (Esri, Redlands, CA). The interactive web version of the analysis is available as a public ArcGIS StoryMap.

### Ethics

This analysis used exclusively publicly available, aggregated, de-identified data at the census-tract level, together with facility-level location data that did not include any patient information. Because the work does not involve identifiable private information about living individuals, it does not meet the regulatory definition of human-subjects research under 45 CFR 46.102(e). Institutional review-board (IRB) oversight was therefore not sought.

## Results

### Distribution of CT imaging infrastructure

We identified 408 CT imaging facilities across the Greater Houston metropolitan area, comprising 108 hospital-based scanners, 90 independent imaging centers, and 210 freestanding emergency facilities with CT. Facilities were markedly clustered in the central urban core and along higher-income corridors radiating from the Texas Medical Center (Figure 1). Hospital-based scanners, independent imaging centers, and freestanding emergency facilities showed broadly similar geographic footprints: all three facility types concentrated disproportionately within the Houston city limits and in affluent inner suburbs, with comparatively sparse coverage in peripheral, peri-urban, and southeastern tracts.

### CT access and poverty

Overlaying CT facility locations on the census-tract poverty choropleth revealed a pronounced negative spatial association between poverty and diagnostic capacity (Figure 2). Tracts with poverty rates above 24%, concentrated on the eastern and southeastern periphery of the city, contained markedly fewer CT scanners than tracts with poverty rates below 10%. Hospital-based and independent scanners were noticeably clustered in less impoverished tracts, revealing deep structural gaps in where advanced diagnostic services are physically placed.

### CT access and uninsured rate

A similar pattern was observed when CT facility locations were overlaid on the census-tract uninsured-rate choropleth (Figure 3). Tracts with fewer than 1% of residents uninsured — largely in the central and near-western parts of the city — contained a dense concentration of CT scanners, whereas tracts with uninsured rates exceeding 16% — predominantly in the eastern and southeastern periphery — contained markedly fewer CT imaging facilities. The pattern persisted across facility type, indicating that freestanding emergency and independent imaging centers did not meaningfully compensate for the undersupply of hospital-based scanners in higher-uninsured communities.

### CT access and median household income

The income choropleth reproduced the same broad gradient (Figure 4). CT scanners clustered disproportionately in dark-green tracts with median household income above $119,300, while purple tracts with median household income below $37,800 — concentrated along the eastern, southeastern, and far-western periphery — exhibited substantially fewer nearby diagnostic options. Middle-income tracts (light green; near the $78,500 national median) occupied an intermediate position, with CT coverage that was better than in low-income tracts but consistently thinner than in the highest-income tracts.

### Convergent geographic pattern across indicators

Taken together, the three socioeconomic overlays (Figures 2–4) describe a single convergent geographic pattern: CT imaging infrastructure in Greater Houston is concentrated in tracts that are simultaneously lower-poverty, higher-income, and better-insured, and is relatively sparse in tracts that are simultaneously higher-poverty, lower-income, and higher-uninsured. In a geographically large and car-dependent city such as Houston, in which many lower-income residents rely on limited public transportation, this distribution represents a substantive barrier to timely diagnostic imaging for the communities most likely to experience downstream morbidity.

## Discussion

In this citywide geospatial analysis of Greater Houston, the distribution of CT imaging infrastructure mirrored — rather than offset — underlying socioeconomic inequality. Across three complementary indicators of neighborhood-level vulnerability (poverty rate, uninsured rate, and median household income), CT facilities clustered consistently in the least vulnerable tracts, and a spatial gap in advanced diagnostic capacity was observed on the eastern and southeastern periphery of the city. The result is a diagnostic-infrastructure pattern in which the communities whose residents are most likely to present late, with advanced disease, or through the emergency department, also face the greatest travel and coverage barriers to obtaining the cross-sectional imaging that drives diagnosis and triage.

These findings are consistent with prior work documenting geographic concentration of advanced diagnostic resources in high-income urban cores [5–7] and with analyses showing that the sheer presence of a major medical complex does not by itself guarantee equitable spatial access [9]. The study extends that literature by characterizing the pattern across multiple facility types simultaneously — hospital-based, independent, and freestanding emergency — and by showing that the alternative facility types did not meaningfully compensate for the undersupply of hospital-based scanners in vulnerable tracts. Similar observations have been reported for other advanced modalities such as MRI and PET [4, 6, 10], and together they suggest a systemic rather than modality-specific pattern of infrastructural inequity.

From a policy perspective, the analysis supports three broad implications. First, health-system planning and certificate-of-need style review processes might incorporate explicit spatial-equity criteria when evaluating proposals for new or expanded imaging capacity, rather than relying on aggregate citywide counts [11]. Second, targeted investment — including mobile CT, community-embedded imaging hubs, and transportation supports — may be warranted in tracts identified as simultaneously high-poverty, high-uninsured, and under-served by existing CT infrastructure [7, 12]. Third, safety-net systems and federally qualified health centers (FQHCs) may be well positioned to partner with hospital-based imaging programs to extend capacity into these tracts [12].

### Strengths and limitations

Strengths of this analysis include its use of three complementary socioeconomic indicators, inclusion of multiple facility types, and use of census tract–level resolution, which is finer than the county- or ZIP-level resolution used in many prior imaging-access analyses. The study has several limitations. First, the facility roster, while triangulated across three sources, may not capture every CT scanner operating in the metro area, particularly newly opened independent centers; however, any such omissions are unlikely to reverse the central geographic pattern. Second, the analysis is cross-sectional and does not capture temporal dynamics such as facility openings or closures. Third, the analysis is descriptive and does not formally model travel time, scanner throughput, or per-capita demand; future extensions should incorporate enhanced two-step floating catchment area methods, drive-time network analysis, and public-transit accessibility [5, 6]. Finally, the analysis does not capture differences in appointment wait times, after-hours availability, or accepted insurance plans, which are important non-spatial dimensions of effective access [7].

## Conclusions

In Greater Houston, the spatial distribution of CT imaging infrastructure mirrors rather than offsets underlying socioeconomic inequality: diagnostic capacity is concentrated in lower-poverty, higher-income, and better-insured tracts, and is comparatively sparse in tracts with higher poverty, lower income, and higher uninsured rates. Citywide spatial analysis renders these inequities visible in ways that individual clinical encounters cannot. These findings suggest that equitable access to advanced diagnostic care requires explicit attention to geography alongside clinical need and that investment strategies, partnerships with safety-net providers, and policies that link imaging-capacity expansion to measurable community need may help to close the observed gap.

## Data Availability

All data analyzed in this study are derived from publicly available sources. American Community Survey (ACS) 5-year estimates are available from the U.S. Census Bureau at https://data.census.gov. Centers for Medicare and Medicaid Services (CMS) provider data are available at https://data.cms.gov. Texas Department of State Health Services (DSHS) facility licensure information is available at https://www.dshs.texas.gov/health-facility-regulation. The aggregated CT facility roster, the joined census tract-level analytic dataset, and the accompanying ArcGIS StoryMap are available from the corresponding author upon reasonable request.

https://data.census.gov

https://data.cms.gov

https://www.dshs.texas.gov/health-facility-regulation

## List of abbreviations

2SFCA: two-step floating catchment area
ACS: American Community Survey
BMC: BioMed Central
CMS: Centers for Medicare & Medicaid Services
CT: computed tomography
DSHS: Texas Department of State Health Services
FQHC: federally qualified health center
GIS: geographic information system
IRB: institutional review board
MPH: Master of Public Health
MRI: magnetic resonance imaging
PET: positron emission tomography
TMC: Texas Medical Center

## Declarations

### Ethics approval and consent to participate

Not applicable. This study used exclusively publicly available, aggregated, de-identified data at the census-tract level and facility-level location data that did not contain any information about identifiable living individuals. The work does not meet the regulatory definition of human-subjects research under 45 CFR 46.102(e); institutional review-board (IRB) oversight was therefore not sought, and informed consent was not applicable.

### Consent for publication

Not applicable.

### Availability of data and materials

The datasets generated and analysed during the current study are derived from publicly available sources. American Community Survey data are available from the U.S. Census Bureau (https://data.census.gov). Centers for Medicare & Medicaid Services provider data are available at https://data.cms.gov. Texas DSHS facility licensure information is available at https://www.dshs.texas.gov/. The aggregated facility roster, the joined tract-level analytic dataset, and the accompanying ArcGIS StoryMap are available from the corresponding author upon reasonable request. CT imaging location data contributed by MD Anderson Cancer Center are available subject to that institution’s data-sharing policies.

### Authors’ contributions

DMO conceived and designed the study, compiled and curated the facility roster, assembled the ACS, CMS, and DSHS data, performed the geocoding and spatial overlay analysis, produced all maps, and drafted and revised the manuscript. The author has read and approved the final manuscript.

## Acknowledgements

The author thanks Dr. Claudia C. Tamara (The University of Texas MD Anderson Cancer Center) for mentorship and guidance throughout this project, and the faculty of the Brown University School of Public Health for their support of the Master of Public Health Practicum. The author also acknowledges the U.S. Census Bureau, the Centers for Medicare & Medicaid Services, the Texas Department of State Health Services, OpenStreetMap contributors, Esri, and the City of Houston GIS program for providing the open data and tooling that made this analysis possible.

